# No One Left Behind: Adaptive Tablet Modalities for Digitally Excluded Emergency Department Patients Design, Implementation, and Social Evidence for an Impairment-First Interface

**DOI:** 10.64898/2026.04.11.26350686

**Authors:** Abir Chowdhury, Atashi Irtiza

**Affiliations:** MSc Engineering — Innovative Communication Technologies and Entrepreneurship Aalborg University, Copenhagen, Denmark; Department of Ophthalmology, Bangladesh Medical University, Dhaka, Bangladesh

**Keywords:** adaptive interface, digital health equity, emergency department, impairment-first design, accessibility, assistive technology, health informatics, social media discourse analysis, patient dignity, alarm fatigue, triage literacy

## Abstract

**Background:** The urgent care departments in Europe face a structural paradox: accelerating digitalisation is accompanied by a patient population that is disproportionately unable to engage with standard digital tools. An internal analysis at the Emergency Department (Akutafdelingen) of Nordsjællands Hospital in Hillerød, Denmark found that 43% of emergency patients struggle with digital solutions — a figure that reflects the predictable composition of acute care populations rather than any individual failing.

**Objective:** This paper presents the design, iterative development, and secondary validation of the ED Adaptive Interface (v5): a prototype adaptive patient terminal developed in response to this challenge. The system operationalises what the author terms impairment-first design — a methodology that treats the most constrained patient experience as the primary design problem and derives the standard experience as a subset. The interface configures itself in under ten seconds via nurse-led setup, adapting across four axes of impairment: visual, motor, speech, and cognitive.

**System:** Version 4 supports five accessibility modes, a heatmap pain assessment grid, a Privacy and Dignity panel, a live workflow tracker with care notifications, structured dual-category help requests, and plain-language medical term definitions across four languages. Version 5, reported here for the first time, introduces a Condition Worsening Escalation button, a Referral Pathway Display, a "Why Am I Waiting?" triage explainer, a Symptom Progression Log, MinSP/Yellow Card Scan simulation, expanded language support (seven languages: English, Danish, Arabic with full RTL layout, Turkish, Romanian, Polish, and Somali), and an expanded ten-item Communication Board. The entire system runs as a single 79-kilobyte HTML file with zero infrastructure requirements.

**Methods:** To base the design on patient-generated evidence, two independent social media threads were subjected to an inductive thematic analysis (Braun and Clarke, 2006) a primary corpus of 83 entries in the Facebook group: Foreigners in Denmark (collected March 2026) and a corroborating corpus in an international community group in the Aarhus region (collected April 2026). All identifiers in both datasets were fully anonymised under GDPR Article 89 research provisions prior to analysis. No participants were contacted. Generative AI tools were used to assist with drafting, writing, and prototype code development in the preparation of this manuscript; all scientific content, data collection, analysis, and conclusions are the sole responsibility of the authors.

**Results:** The first discourse corpus produced five major themes in relation to the five general problem areas that the prototype was intended to cover: system navigation and triage literacy gaps (31 entries); language and cultural barriers (6 entries); communication failures during care (5 entries); staff overload and capacity constraints (8 entries); and pain and severity assessment failures (14 entries). The supportive dataset supported all five themes on its own and presented two new themes: the different treatment of international patients and medical gaslighting as a long-term trend of patient advocacy failure. One of the major structural discoveries the five most-liked comments were critical of the original poster being self-referring to the ED when she had in fact been explicitly triaged to receive 1813 telephone referral to the ED directly inspired the Referral Pathway Display and Why Am I Waiting? features in v5.

**Conclusions:** The convergence of design rationale and independent social evidence across all five problem categories suggests that impairment-first design is not a niche accessibility concern but a structural approach to healthcare interface quality. The prototype is ready for a structured clinical pilot using the System Usability Scale (SUS) and semi-structured staff interviews. The long-term roadmap includes full MinSP integration, hospital PMS connectivity, and clinical validation.

## 1. Introduction

Healthcare systems across Europe face a structural paradox. The same demographic groups most likely to require emergency care — older adults, patients with chronic conditions, recent migrants, and individuals with cognitive or physical impairments — are also those least equipped to navigate the digital interfaces through which care is increasingly delivered. As healthcare digitalisation accelerates, this asymmetry does not resolve itself; it compounds.

At Nordsjællands Hospital’s Emergency Department (Akutafdelingen) in Hillerød, Denmark, the tension between digitalisation and inclusion arrived as a concrete institutional problem. A hospital-led internal analysis determined that 43% of emergency patients struggle with digital solutions. The Emergency Department sees an unpredictable cross-section of the population: an 85-year-old arriving with acute confusion sits alongside a 30-year-old with a sports injury; a recent refugee with no Danish shares a waiting area with a long-term resident who has no smartphone and no prior encounter with digital healthcare tools. Generic tablet interfaces, calibrated for a digitally fluent adult in a state of baseline calm, exclude nearly half of this population before a single clinician has entered the room.

The solution concept described in this paper originated during the OpenInnovation 2026 Health challenge — a cross-university student innovation sprint co-organised by DTU Skylab, Copenhagen School of Entrepreneurship (CBS), and KU Actory, with Nordsjællands Hospital as the case partner — but the prototype, methodology, and all work reported here were developed independently after that event. The ED Adaptive Interface is a working prototype that configures itself in under ten seconds to serve patients across the full spectrum of visual, motor, speech, and cognitive impairment, in their own language, with zero infrastructure requirements.

The design philosophy at the core of the prototype — *impairment-first design* — is the paper’s central theoretical contribution. Rather than building an interface for a typical user and retrofitting accessibility as a compliance layer, the design began with the most constrained patient imaginable: one who cannot see, cannot touch the screen, cannot speak, and cannot read Danish. Every design decision was tested against this patient’s needs before any other consideration. The result is an interface that, by solving the hardest problem first, also serves every other patient better.

This paper makes four contributions. First, it documents the architecture and full feature set of v4 of the ED Adaptive Interface. Second, it reports v5 — a set of evidence-driven upgrades developed following secondary validation through social media discourse analysis — for the first time. Third, it formalises the impairment-first methodology as a transferable framework for healthcare interface design. Fourth, it presents an inductive thematic analysis of 83 entries from a public social media forum as a post-hoc validation source — an approach that, to the author’s knowledge, has not previously been applied in this form in health informatics research.

The prototype is technically unremarkable: a single HTML file, 79 kilobytes, running on any modern browser without installation or network connection. The contribution is not the technology. It is the thinking that produced it, and the evidence that the problems it addresses are real, widespread, and structurally predictable.

## 2. Background and Related Work

### 2.1 Digital Exclusion in Healthcare Settings

The relationship between digitalisation and equity in healthcare is well-established in the literature. Digital health interventions consistently assume levels of literacy, language proficiency, motor dexterity, and cognitive capacity that many patients — particularly those most likely to require emergency care — cannot reliably provide [1, 2]. This is not a minor oversight. Older adults, migrants, patients with physical disabilities, and those experiencing acute psychological distress face compounding barriers that standard interfaces are simply not built to accommodate [3].

In the Danish context, two pressures converge. The country’s commitment to a digital-first public sector has been ambitious and rapid, with electronic patient records, online appointment systems, and digital prescription management all migrating to digital-only delivery within a comparatively short timeframe. Meanwhile, the Emergency Department population is structurally heterogeneous. Eurostat data indicate that between 35 and 50 per cent of adults aged 65 and over in EU member states report significant difficulty with digital devices [4], a proportion that rises sharply in acute care settings where IV lines restrict hand movement, sedation reduces response precision, pain degrades concentration, and anxiety compounds every other barrier.

The 43% figure cited by Nordsjællands Hospital is the predictable outcome of deploying technology designed for baseline conditions in a setting where baseline conditions are the exception. Designing around it is a clinical necessity, not an optional accessibility accommodation.

### 2.2 Accessibility Frameworks and Adaptive Interface Design

The human-computer interaction literature offers well-developed frameworks for accessible design, most prominently the Web Content Accessibility Guidelines (WCAG 2.1) [5] and the WHO’s global assistive technology standards [6]. However, most digital health implementations treat these frameworks as compliance checklists applied after the primary interface is built — a set of retrofitted accommodations for an assumed minority, rather than foundational design constraints.

Universally designed interfaces — those built from the outset for the widest possible range of users — consistently outperform retrofitted accessible variants across usability and satisfaction metrics [7]. The curb-cut effect, originally observed in built environments, is well-documented in digital contexts: features introduced for motor-impaired users (keyboard navigation, large tap targets, high contrast modes) reliably improve the experience for all users [8]. Switch-access devices, sip-and-puff technology, and augmentative and alternative communication (AAC) boards are established assistive tools with strong clinical evidence bases [9], yet they remain largely absent from hospital-issued patient-facing interfaces. The ED Adaptive Interface integrates compatibility with these devices natively through its Auto-Scanning Focus mode, at no additional hardware cost.

### 2.3 Alarm Fatigue and Structured Clinical Communication

Emergency departments present a distinctive communication environment. Patients are frightened, frequently in pain, and often unable to articulate their needs clearly. Nursing staff operate under persistent time pressure and high cognitive load. The combination creates conditions for what the clinical literature terms alarm fatigue: the desensitisation to alert signals that develops when the same notification mechanism is used regardless of clinical urgency [10, 11]. When a request for a blanket generates the same alert as a report of chest pain, response thresholds rise over time and genuinely urgent signals risk being missed. Structured communication approaches separating non-urgent comfort needs from clinical urgency have been proposed in nursing informatics [11], but their implementation in patient-facing interfaces has been limited. This gap directly informed the division of all patient help requests into two structurally distinct categories — Comfort and Medical — each generating a different signal type to the care team.

### 2.4 Language Barriers and Triage Safety

Language discordance between patients and providers is a documented patient safety risk in emergency settings. Flores’s systematic review [12] identified language barriers as contributing to adverse events, reduced symptom accuracy at triage, and lower patient satisfaction. In Danish emergency departments, conservative estimates suggest that over 20% of patients speak Danish as a second language. Existing solutions — telephone interpreters, printed multilingual handouts — share a common limitation: they depend on staff initiative, introduce logistical delay, and are unavailable at first patient contact. An interface that delivers every element of the patient experience in the patient’s own language automatically from the moment of nurse setup represents a qualitatively different approach.

### 2.5 Pain Assessment in Emergency Populations

Standard pain assessment tools — the Visual Analogue Scale (VAS) and Numeric Rating Scale (NRS) — were validated primarily in research populations. Their performance degrades in patients with limited literacy, language barriers, motor impairment, or cognitive distress, precisely the populations most common in emergency settings [13]. Multi-modal pain assessment tools presenting simultaneous numerical, colour, and facial expression cues have demonstrated superior reliability in these populations. The heatmap pain grid in the ED Adaptive Interface combines these three cue types with large tap targets compatible with tremor, IV lines, and agitation.

### 2.6 Health Literacy and Patient Engagement

Health literacy is a well-established predictor of healthcare outcomes. Paasche-Orlow and Wolf [14] identified low health literacy as a causal pathway to worse clinical outcomes, lower medication adherence, and higher emergency readmission rates. In an emergency department context, patients frequently encounter medical terminology without explanation, compounding anxiety and reducing cooperation. Plain-language definitions of every medical term appearing in the ED Adaptive Interface, available in all supported languages, represent a low-cost, high-impact response to this documented problem.

## 3. Design Methodology: Impairment-First Design

### 3.1 The Core Principle

The central design principle of the ED Adaptive Interface is impairment-first design. The principle is deceptively simple: identify the most constrained user scenario and solve that problem first. Every subsequent design decision is evaluated against this primary constraint before any other consideration. Typically-abled, digitally literate, linguistically fluent patients are treated not as the default user but as a beneficiary subset — they receive the same interface, which turns out to be better for them too.

This represents a deliberate inversion of the conventional product design hierarchy. In most digital product development, accessibility features are added after the core product is built for a typical user. Impairment-first design inverts this entirely: every feature is built from the ground up to serve the most constrained user, and the standard experience is derived from that. In the words of the design philosophy that guided every decision:

> *"We don’t ask the patient to learn our technology; our technology learns the patient."*

This is not merely a tagline. It describes a specific design protocol. When any feature was under consideration, the first question asked was: can a patient who is blind, paralysed, mute, confused, and reads no Danish use this? If not, the design was rejected or modified until that patient could engage with it. Only then was the feature evaluated against any other criterion.

### 3.2 Defining the Four Impairment Axes

Four dimensions were identified along which patients may experience reduced digital interaction capacity in an emergency context. These are not clinical diagnoses but functional states — the actual capabilities that determine whether a patient can engage with a tablet during an ED stay:

#### Visual

Reduced ability to perceive screen content, due to low vision, blindness, sedation, poor lighting, anxiety, or the absence of reading glasses.

#### Motor

Reduced ability to touch or manipulate a touchscreen, due to tremors, IV lines, post-stroke weakness, paralysis, cerebral palsy, sip-and-puff dependency, or post-operative restriction.

#### Speech

Reduced or absent spoken communication capacity, due to tracheotomy, stroke, severe pain, acute respiratory distress, laryngectomy, or psychological trauma.

#### Cognitive

Reduced ability to navigate multi-step digital interfaces, due to dementia, acute confusion, sedation, high anxiety, acquired brain injury, or simple unfamiliarity with digital tools.

These axes are not mutually exclusive and must be treated as independently composable. The nurse setup interface allows any combination of accessibility flags to be set simultaneously.

### 3.3 The Nurse-Led Configuration Approach

A critical design constraint was time. An admitting nurse typically has seconds, not minutes, to configure any patient-facing tool. Self-configuration approaches are inappropriate for populations that are, by definition, struggling with the technology. The solution was a dual-tab nurse setup interface. The first tab asks what the patient can do (abilities); the second asks what barriers they face (disabilities). Leading with capability rather than limitation produces a more complete and more dignified patient profile. The complete configuration requires fewer than ten seconds, after which the system transforms automatically to the configured mode.

### 3.4 Iterative Development Process

The prototype was developed iteratively, progressing from v1 through v4 over the course of the OpenInnovation 2026 challenge, with each version documented in external-facing specifications — the Feature Guide and Jury Q&A documents — that maintained design coherence. Version 5, reported in this paper, was developed independently after the challenge period, driven directly by findings from the social media discourse analysis described in Section 5. The v5 features are not additions motivated by developer preference but upgrades traceable to specific themes in patient-generated discourse data.

## 4. System Architecture and Feature Implementation

### 4.1 Technical Architecture

The ED Adaptive Interface is implemented as a single HTML file using React 18.2 (loaded via CDN) and Tailwind CSS for styling. The compiled application is approximately 79 kilobytes — smaller than the average web page image. It runs fully offline in any modern browser without installation, server infrastructure, or network connectivity. There is no database, no user account system, no app store, and no IT department involvement required. The file can be transferred to a hospital tablet via USB drive and opened directly.

This zero-infrastructure architecture was a deliberate design constraint, not a technical limitation. Hospital IT environments are typically slow to onboard new software, requiring security audits and presenting significant barriers to rapid deployment. A self-contained HTML file bypasses all of these barriers: any ward with a browser can run the prototype immediately. The absence of server-side data persistence is acceptable at prototype stage and is addressable through future integration with the hospital’s patient management system (PMS).

### 4.2 Nurse Setup Interface

The nurse setup screen is divided into two tabs: Abilities (what the patient can do) and Disabilities (barriers they face). Language selection is a global toggle affecting every string in the entire interface. Accessibility mode selection is checkable in any combination. From v5, the nurse also selects the patient’s referral source — 1813 telephone triage, self-referral, ambulance, or GP — which then appears as a persistent badge on every patient-facing screen for the duration of the visit.

A MinSP / Yellow Card Scan button in v5 simulates the integration concept for the long-term roadmap. The nurse taps "Scan Now," an animated spinner plays, and the patient profile is pre-populated with placeholder data. This is a proof-of-concept demonstration, not a live integration; its purpose is to show stakeholders what the completed integration pathway would look like in practice.

### 4.3 Accessibility Modes

Five accessibility modes transform the patient dashboard automatically based on the nurse’s configuration. Each mode is independently activatable and composable with others. Table 1 summarises the five modes, their target impairment axes, patient experience, and technical implementation.

**Table 1.** Accessibility modes in the ED Adaptive Interface v4/v5.

| Mode | Activates For | Patient Experience | Technical Implementation |
| --- | --- | --- | --- |
| High Contrast | Visual impairment | Blue/Orange palette (WCAG AA, colorblind-safe); 50% larger text; enlarged tap targets | Conditional Tailwind class assignment on all components |
| Talk-Aloud | Blind / low vision | Every interactive element reads its label aloud when focused; speaker icon visible on every button | Browser SpeechSynthesis API; event-driven utterance queue |
| Auto-Scanning Focus | Motor impairment | Glowing border cycles through buttons every 3 seconds; any keypress or external switch selects highlighted element | useEffect with setInterval; keydown event listener; CSS keyframe animation |
| Voice Command Mode | Speech-limited (residual) | Pulsing microphone + voice-tag overlays showing spoken commands (e.g., "Say 'Help'") | Absolutely positioned overlay elements; SpeechRecognition API (graceful degradation) |
| Cognitive Workflow Tracker | Cognitive impairment | Linear five-stage tracker (Triage → Tests → Doctor → Treatment → Discharge) dominates home screen; plain-language explanation per stage | State-driven rendering with animated progress component |

The Blue/Orange high-contrast palette warrants specific explanation. Most high-contrast implementations use black-and-white, which fails for the most common forms of colour vision deficiency (red-green deficiency, affecting approximately 8% of males of Northern European descent). Blue-orange combinations are discriminable by approximately 99% of the population, including those with deuteranopia and protanopia, while still achieving the luminance contrast ratios required by WCAG 2.1 Level AA [5].

The Auto-Scanning Focus mode is fully compatible with external switch devices, including sip-and-puff systems used by patients with severe motor impairment (ALS, high spinal injury, locked-in syndrome). A glowing border cycles through interactive elements on a three-second interval; any external keypress event — from a keyboard, a Bluetooth switch, or a sip-and-puff device — triggers selection of the currently highlighted element. The interface is fully operable by patients who have zero direct contact with the touchscreen, at no additional hardware cost.

### 4.4 Clinical Communication Features

#### 4.4.1 The Divided Help Request System

All patient help requests are divided into two structurally distinct categories. Comfort requests water, blanket, temperature adjustment, privacy, dimmed lights — are non-urgent and generate a low-priority signal. Medical requests — pain, difficulty breathing, dizziness, chest pain — are urgent and generate an escalated signal requiring prompt attention. The clinical rationale is alarm fatigue prevention [10]: when every request generates the same alert regardless of urgency, response thresholds rise over time and critical signals risk being missed.

#### 4.4.2 Heatmap Pain Assessment Grid

The standard Visual Analogue Scale requires fine motor control and presents a single positional cue that may not carry consistent meaning across language and cultural contexts. The ED Adaptive Interface replaces this with a grid of eleven large tap targets (0–10) providing three simultaneous cues per rating: a colour (green to red gradient), a number, and an emoji icon. The targets are sized to be reliably activatable by patients with IV lines, tremors, or motor agitation. This approach is consistent with recommendations in the pain assessment literature for multi-modal representations in linguistically and cognitively diverse populations [13].

#### 4.4.3 Privacy and Dignity Panel

The Privacy and Dignity Panel provides one-tap access to six requests without requiring speech, explanation, or waiting for a nurse to notice: Close Curtain, Cover Me, Lower Lights, Need Privacy, Call Family, and Feel Unsafe. The last of these is a discreet escalation mechanism — a silent alert to staff that the patient feels unsafe — without requiring the patient to speak, raise their hand, or attract attention within the department. To the author’s knowledge, this feature is absent from all existing commercial ED patient terminals.

#### 4.4.4 Live Workflow Tracker and Care Notifications

A five-stage workflow tracker (Triage → Tests → Doctor → Treatment → Discharge) provides patients with a persistent visual representation of where they are in the care process. At each stage transition, a banner notification appears in the patient’s own language explaining what just happened and what comes next, written in plain language. Proactively answering "What is going on?" before the patient has to ask eliminates a category of nursing interruption that consumes significant time in high-volume ED settings.

#### 4.4.5 Communication Board

The Communication Board provides tap-to-talk access to frequently needed phrases for non-verbal or speech-impaired patients. Version 4 included six items: Thirsty, Bathroom, Cold, Yes, No, and More Pain. Version 5 expanded this to ten items, adding Scared, Too Loud, Allergies!, and I Need Help — additions directly motivated by entries in the social media discourse corpus describing patients who had no means to communicate emotional distress or urgency escalation. When the speech-impaired mode is active, the Communication Board is promoted to the home screen.

#### 4.4.6 Plain-Language Medical Term Definitions

Every medical term appearing in the interface — Triage, IV Line, Vital Signs, ECG, and others has an expandable plain-language definition available in all supported languages. Health literacy research consistently shows that patients who understand their clinical situation are more cooperative care partners and report better satisfaction outcomes [14]. Providing this information passively at the point of encounter reduces both cognitive burden on the patient and the frequency of explanatory interruptions to nursing staff.

### 4.5 Version 5 New Features

Version 5 introduced seven substantive upgrades, all directly traceable to themes identified in the social media discourse analysis reported in Section 5. Table 2 summarises these additions.

**Table 2.**
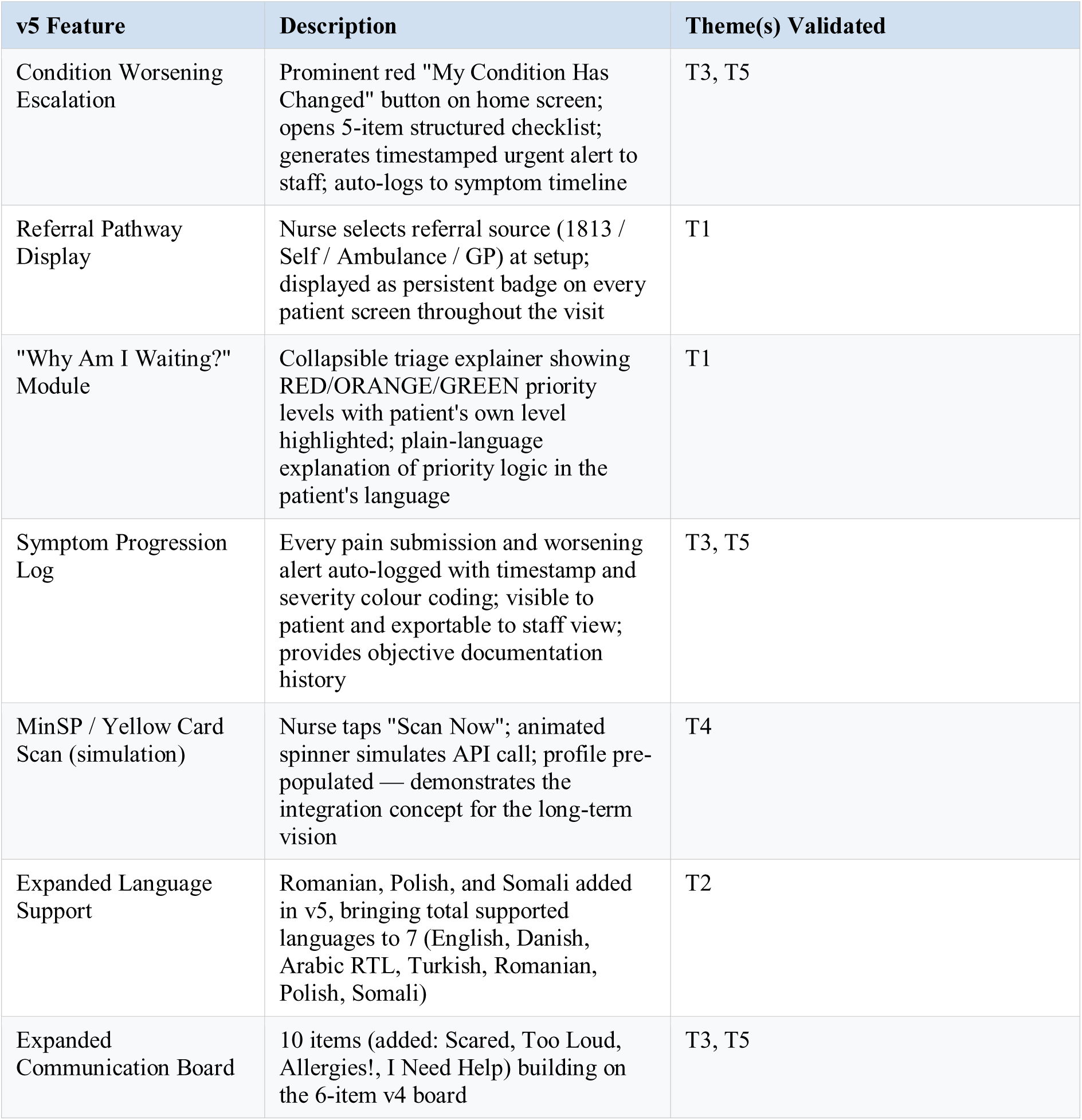
Version 5 additions to the ED Adaptive Interface, mapped to discourse themes.

| v5 Feature | Description | Theme(s) Validated |
| --- | --- | --- |
| Condition Worsening Escalation | Prominent red "My Condition Has Changed" button on home screen; opens 5-item structured checklist; generates timestamped urgent alert to staff; auto-logs to symptom timeline | T3, T5 |
| Referral Pathway Display | Nurse selects referral source (1813 / Self / Ambulance / GP) at setup; displayed as persistent badge on every patient screen throughout the visit | T1 |
| "Why Am I Waiting?" Module | Collapsible triage explainer showing RED/ORANGE/GREEN priority levels with patient's own level highlighted; plain-language explanation of priority logic in the patient's language | T1 |
| Symptom Progression Log | Every pain submission and worsening alert auto-logged with timestamp and severity colour coding; visible to patient and exportable to staff view; provides objective documentation history | T3, T5 |
| MinSP / Yellow Card Scan (simulation) | Nurse taps "Scan Now"; animated spinner simulates API call; profile pre-populated — demonstrates the integration concept for the long-term vision | T4 |
| Expanded Language Support | Romanian, Polish, and Somali added in v5, bringing total supported languages to 7 (English, Danish, Arabic RTL, Turkish, Romanian, Polish, Somali) | T2 |
| Expanded Communication Board | 10 items (added: Scared, Too Loud, Allergies!, I Need Help) building on the 6-item v4 board | T3, T5 |

**Table 3.** Discourse themes and corresponding interface features.

| Theme | Entries (n) | Core Finding | Interface Response |
| --- | --- | --- | --- |
| T1 | 31 | Pervasive confusion about triage priority, waiting logic, and when ED attendance is appropriate; fellow patients misidentified 1813-approved attendances as self-referrals | Why Am I Waiting? module; Referral Pathway Display; Workflow Tracker |
| T2 | 6 | Language and cultural mismatch between patients and staff; secondary social dismissal of linguistically diverse patients | 7-language support (v5); automatic language-matched notifications |
| T3 | 5 | Patients left without updates or communication for extended periods after initial treatment, including one case of patient self-discharge | Care notifications; Condition Worsening Escalation; Symptom Progression Log |
| T4 | 8 | Systemic understaffing attributed as root cause of clinical oversights; staff too stretched to provide adequate communication | Divided Help System; Workflow Tracker (reduces status-update interruptions) |
| T5 | 14 | Pain and symptom severity perceived as dismissed or disbelieved; patients lacked means to communicate deterioration credibly | Heatmap Pain Grid; Symptom Progression Log; Condition Worsening Escalation |

Two v5 features deserve particular elaboration. The Condition Worsening Escalation button addresses the most serious failure mode in the discourse corpus: patients whose clinical deterioration was not communicated to staff, either because they could not attract attention or because their self-reports of worsening were not taken seriously. The button is prominently placed on the home screen, coloured red, and opens a structured five-item checklist before generating a timestamped alert. The structured format reduces false activations and provides staff with specific clinical information before they enter the room.

The "Why Am I Waiting?" module addresses Theme T1 directly. It presents a collapsible explainer showing the triage priority system (RED/ORANGE/GREEN levels) with the patient’s own assigned level highlighted, alongside the referral source badge set by the nurse at admission. A patient who arrived with 1813 approval can see, on their own screen, that they arrived via the telephone triage service and that their priority level is ORANGE. This does not change the wait time, but it fundamentally changes the information environment in which the patient waits — and, as the discourse corpus demonstrates, the absence of this information generates significant distress, confusion, and unjustified social sanction from other patients who assume self-referral.

## 5. Social Media Discourse as Post-Hoc Design Validation

### 5.1 Rationale

The design decisions described in Section 4 were grounded in published accessibility standards, alarm fatigue literature, pain assessment research, and health literacy evidence. But institutional research and academic literature have a well-documented limitation: they tend to capture what healthcare systems choose to measure and report. The frustration of waiting four hours without an update, of having pain dismissed, of not understanding why a patient was lower priority than an ambulance arrival — this texture of lived emergency care experience is underrepresented in formal patient satisfaction instruments. It is, however, articulated in detail in the unsolicited testimony of patients sharing experiences with peers.

A secondary evidence source was therefore sought: unsolicited, peer-generated patient testimony from a public online forum. The goal was not to validate individual anecdotes as statistically representative findings, but to test whether the categories of patient frustration the design addressed were recogniz able in organic patient speech, and whether there were important categories that had been missed.

### 5.2 Data Source and Collection

The primary dataset consists of 83 entries — one original post (OP) and 82 comments — from a thread in the Facebook group “Foreigners in Denmark,” which were gathered on 14 March 2026. The triggering post described three consecutive negative interactions with Danish emergency healthcare: an approved Emergency Department visit that resulted in a four-hour wait without seeing a doctor; a late-night allergic reaction managed via telephone triage; and a clinic scheduling error. The post attracted responses from over sixty distinct commenters over approximately one week. A second, independently collected thread was identified in April 2026 in a Facebook group serving the international community in the Aarhus region of Denmark, described in full in Section 5.9.

This source offers several methodological advantages over formal patient satisfaction instruments. The content was entirely unsolicited, as participants responded to a peer’s lived experience rather than a research prompt, which substantially reduced social desirability bias. The group’s membership (international residents of Denmark) corresponds closely to one of the most linguistically and culturally excluded patient populations in Danish emergency care. The thread captured a genuine, multi-perspective debate including both strongly critical and strongly positive voices.

The thread was archived in full before analysis. All identifiable information was removed and replaced with reference codes (C01–C81 for commenters, OP for the original poster) prior to any coding or analysis. For the corroborating Aarhus dataset, the same anonymisation procedure was applied independently, with separate reference codes assigned. No participant in either dataset was contacted. The data ethics framework governing both analyses is described in the Data Ethics Statement at the end of this paper.

### 5.3 Analytical Method

Inductive thematic analysis was applied following the six-phase framework described by Braun and Clarke [15]: familiarisation with the data; initial code generation; theme searching; theme review; theme definition and naming; and reporting. The analytical process was inductive — themes were generated from the data rather than applied from pre-existing categories — though the emergent themes were subsequently mapped to design features for validation purposes.

Ten themes were identified across the primary 83-entry corpus; the corroborating dataset (Section 5.9) was analysed separately using the same method and produced two additional themes beyond those found in the primary corpus. Five themes from the primary corpus correspond directly to features of the ED Adaptive Interface and are reported in detail in Sections 5.4–5.8 below. The remaining five primary themes — comparative international perspectives on emergency care, debates about the semantics of tax-funded healthcare, GP quality variability, private healthcare alternatives in Denmark, and contextual/demographic variation in experience — are documented in the research archive but fall outside the scope of this paper’s validation analysis.

### 5.4 Theme 1 — System Navigation and Triage Literacy Gaps (n = 31)

The most prevalent theme, present in 31 of 83 entries, concerns patients’ confusion about the correct care pathway — when emergency attendance is appropriate, what the 1813 telephone triage service does, how triage priority is determined, and why waiting times vary so dramatically.

A structural pattern in the high-engagement responses is particularly revealing. The five most highly-liked comments in the entire thread (peak engagement: 243 reactions) all criticised the original poster for "going to the emergency room for a flu," assuming she had self-referred. She had not: she had received explicit 1813 approval, a fact she stated clearly but which multiple high-engagement commenters either did not read or discounted. The comment with the single highest engagement in the thread was therefore factually incorrect about the central fact it was criticising.

This reveals that the information gap about how triage and referral work operates not only between patients and the healthcare system, but within the public understanding of how the system works more broadly. The Referral Pathway Display and Why Am I Waiting? module in v5 address this directly by surfacing the patient’s referral source and triage level on their own screen from the moment of admission.

> *"In the emergency room they have also ambulances bringing patients, so even though you don’t see anyone being taken in, there might be more urgent ones coming. They do triage in emergencies and if your symptoms are not life threatening you need to wait for your turn, even if you are welcomed." — C11 (anonymised)*

### 5.5 Theme 2 — Language and Cultural Barriers (n = 6)

Six entries explicitly named language as a root cause or significant contributing factor to miscommunication. One commenter described the complaints as arising from "a mismatch of expectations and miscommunication" in situations where "there is a language barrier that there isn’t time to deal with." Another directly asked the original poster whether her Danish was sufficient to have correctly understood the appointment time — a question that exemplifies the secondary social harm of language barriers: the deflection of systemic failure onto individual linguistic inadequacy. The seven-language interface, requiring no staff initiative to activate and producing no additional logistical delay, represents a structural response to this structural problem.

### 5.6 Theme 3 — Communication Failures During Care (n = 5)

Five entries described patients who had received initial clinical care and were then left without information, communication, or staff contact for extended periods. The most detailed account described a patient experiencing anaphylaxis who received an adrenaline drip and was left unattended for five hours with no nurse check-in and no information about expected timeline. The patient ultimately self-discharged. These are information failures, not primarily clinical failures. A patient who has been told "we will check on you in approximately ninety minutes" is in a qualitatively different psychological state from a patient who has been waiting ninety minutes in silence. The live workflow tracker, care notifications, and v5 Condition Worsening Escalation button are direct responses to this theme.

### 5.7 Theme 4 — Staff Overload and Systemic Capacity Constraints (n = 8)

Eight entries attributed failures to systemic understaffing rather than individual clinical error. One long-term Danish resident described the system as "breaking down" because doctors and nurses are "too busy and can’t cope." This theme does not validate any specific feature of the prototype — structural understaffing cannot be solved with an HTML file — but it contextualises every feature aimed at reducing nursing workload. An interface that generates structured, categorised requests rather than undifferentiated alerts does not create additional nursing capacity, but it does mean that the nurses who are present spend less time on information-provision tasks a well-designed interface can handle.

### 5.8 Theme 5 — Pain and Severity Assessment Failures (n = 14)

The second most prevalent theme, present in 14 entries, concerns patients who felt their pain or symptom severity was dismissed or under-assessed. Accounts included patients told to "take paracetamol and wait," patients whose self-reported severity was visibly discounted by triage staff, and one account in which a respiratory event was dismissed as "just anxiety" before the situation escalated. The underlying problem, at least in part, is a communication problem. Patients unable to articulate severity clearly, in a state of acute distress, in a language that may not be their own, are structurally disadvantaged. The heatmap pain grid provides a means of communicating severity robust to speech limitation, motor impairment, and linguistic difference. The Symptom Progression Log provides an objective timestamped record of self-reported pain scores — patient-generated evidence harder to dismiss than a single verbal report made during a brief triage encounter.

### 5.9 Corroborating Evidence: Second Discourse Dataset

#### 5.9.1 Data Source

In April 2026 a second independent discourse thread was gathered on a Facebook group that serves the international resident community in the Aarhus region of Denmark. The triggering post narrated about a severe medical crisis that had been faced by a close friend of the post writer about two to three weeks ago. The post was posted directly to create awareness and it received over 100 comments prior to being archived. All identifiers were completely anonymised in the same GDPR Article 89 provisions of the primary dataset. The provisions of data ethics as outlined in the Data Ethics Statement at the end of this paper are applicable to this dataset in their entirety.

#### 5.9.2 Critical Incident: Delayed Appendicitis Diagnosis

The main event narrated in the triggering post is the most clinically serious one that was observed in both discourse datasets. The patient was brought to the emergency service of the Skejby Hospital through the 1813 telephone triage line reporting of severe abdominal pain. The call was dismissed and the triage operator recommended paracetamol. The patient made another call when the pain was increasing and demanded to be attended to and was finally admitted but upon examination, she reported that nothing was wrong. She received a room that lacked a decent bed (as she was informed it was an examination room that had a temporary assessment surface) and medication that failed to alleviate the pain.

During the next three days, or between Tuesday evening and the morning of Friday, the patient repeatedly reported the progression of pain to employees. No additional diagnostic test was done. The patient made a call on Friday morning, three nights later, to 112, who was inside the emergency department, and complained of a possible life-threatening pain, and no diagnosis was made. The national emergency operator noticed that the call was made within a hospital and questioned why. The patient said that she required urgent assistance and no one was attending to her. The emergency personnel liaised with a doctor on duty.

A scan was then requested and found out that there was severe appendicitis and ruptured appendix. Emergency operation was carried out. The Update 1 note on the post indicated that the appendix had already burst by the time the patient was operated on, and that the patient was traumatised, nervous and in shock during the interview. The attending physician who was in charge of the patient after the operation inquired the patient why she had never informed the vagtlaege of her symptoms. She had been trying to communicate them over three days.

There are three v5 features that are directly validated by this incident. The Condition Worsening Escalation button offers a formalized and time-stamped system of patient reporting of clinical deterioration without the use of verbal self-advocacy that can be overlooked. The Symptom Progression Log will provide an objective, chronological record of patient-reported symptom severity that will be with the patient during the visit and will be visible to all clinical personnel - a record that would have been present in this case since Tuesday evening. The context of the information offered by the Why Am I Waiting? module and Referral Pathway Display is that the patients and the staff know the foundation of the attendance, which minimizes the possibility of a 1813-approved, truly acute patient being handled as a low-priority self-referral.

#### 5.9.3 Theme 6: Differential Treatment of International Patients

The comments thread revealed a theme that was not present in the primary data in its explicit form: the different treatment of the international patients as a specific pattern, and not an incidental effect of the language barriers per se. Several commentators, including Danish citizens, reported that there was a tendency where patients with foreign accents, non-Danish names, or poor fluency in the Danish language were more likely to be dismissed. One of the commenters mentioned that he once had a phone call that was ended after he asked to talk in English and the vagtlaege service denied the request. Another one was informed that the system was good with Danes in mind and that other standards were applied to others. One commentator, who described himself as an international MD with five years of experience in Denmark, said that during all the time they worked in the country, no doctor had ever conducted any physical examination on them, such as palpation of the abdomen or listening to the lungs, in a situation where such an examination would be considered a normal clinical procedure in other countries.

This theme expands Theme T2 in the main data set. The structural characteristic of multilingual emergency care is language barriers and cultural mismatch, which have been thoroughly reported in the literature. The differential treatment recorded here, however, transcends a communication difficulty into a pattern of less clinical thoroughness that is associated with patient origin. The seven language support of the ED Adaptive Interface supports the communication dimension; the display of the triage level and referral source to the patient - introduced in v5 supports the information asymmetry dimension. The structural aspect of the differential clinical treatment cannot be addressed in an interface prototype, yet it can be addressed in terms of recording it as a patient safety issue that needs to be addressed systemically.

*"Danish healthcare is the worst nightmare. The system belittles patients, they keep telling us to drink some water and take ibuprofen. Doctors take home big paycheck and lounging like a 5-9 job. I’m sorry but you’re a doctor!!" — Anonymized commenter, International Community Group, April 2026*

#### 5.9.4 Theme 7: Medical Gaslighting and Patient Advocacy Failure

One trend that can be observed in the second dataset that was not highly coded in the primary corpus relates to what several commentators expressly referred to as medical gaslighting, or the systematic rejection of patient-reported symptoms to the extent that patients doubted their own understanding of their health. A commenter said that she was kind and submissive until she was compelled to work hard, and then the staff would become responsive; she said this was the only sure way of getting appropriate care. Another summed up the trend, thus: ignore you until you are so sick you cannot be ignored, then blame you because you got so sick. One of them reported taking months to work between a GP and the emergency room until a cough was diagnosed as pneumonia, by which time it had caused permanent asthma.

This theme is differentiated by Theme T5 (pain and severity assessment failure) in that it is not just the rejection of a single report but a long-term tendency in which patient self-advocacy is systematically suppressed in a variety of contacts. The Condition Worsening Escalation option in v5 is a partial solution: it provides patients with a structured, clinically formatted option whereby they can report and record deterioration in a format that is less easily disregarded than verbal self-reports. By providing a visible and time-stamped log of all pain submissions at the visit, the Symptom Progression Log offers a kind of contemporaneous documentation that, in addition to patient advocacy, minimizes the loss of information that facilitates the continuation of the pattern.

#### 5.9.5 Complaint System Efficacy and the Accountability Gap

One of the repeated sub-themes in the second dataset remarks was the low effectiveness of formal complaint mechanisms. One of the commenters, a Danish citizen who had filed a formal complaint with Styrelsen for Patientklager (the Danish Patient Complaints Board) complained that it took two years before his complaint was dealt with and that the doctors had been found to have dealt perfectly. A number of other commenters recommended not to file complaints on the basis that the system defends itself. The latter perception aligns with the research on patient advocacy that has demonstrated that the complaint systems in healthcare are more likely to favour institutional actors and may cause a major psychological cost to patients who are already in a state of health-related distress.

This sub-theme is not an incentive to a particular new interface feature in the existing prototype but is applicable to the overall research context. When combined with the hospital patient management systems in future development, the Symptom Progression Log in v5 would generate contemporaneous, patient-generated clinical records that would potentially be used to support submissions of complaints with objective evidence of timestamps. The design of sustainable documentation based on internalization of the care encounter, i.e. independent of patient recollection or staff documentation, is a long-term design contribution to the accountability infrastructure of emergency care.

## 6. Discussion

### 6.1 Impairment-First Design as a General Framework

The impairment-first principle is not specific to emergency department tablets or to the Danish healthcare context. The core claim — that designing for maximal constraint produces better outcomes for all users — is consistent with a substantial body of evidence in universal design research [7, 8]. Wheelchair ramps benefit parents with pushchairs, delivery workers, and cyclists. Closed captions benefit viewers in noisy environments. Automatic door openers benefit patients with trolleys and full hands. The curb-cut effect is a structural consequence of solving hard problems thoroughly.

In digital health, investment in accessibility for the most constrained users is not a cost imposed by a niche use case but a structural quality investment that improves the product for every user. Large tap targets make interfaces easier to use for anyone in a stressful, time-pressured environment. Plain-language definitions reduce anxiety for all patients, not only those with limited health literacy. Healthcare institutions that ask "which interface works for the most patients?" systematically underinvest relative to those that ask "which interface works for the patient who arrived alone, in pain, unable to speak, unable to read Danish, and currently at the limits of their cognitive capacity?" The latter question produces better software.

### 6.2 The Social Media Discourse Method: Advantages and Limitations

The use of public social media discourse as a post-hoc design validation source is unusual in health informatics. Its genuine methodological advantages — unsolicited content, unfiltered emotional register, multi-perspective structure — cannot be replicated in formal patient satisfaction surveys. However, several limitations must be acknowledged. The "Foreigners in Denmark" group is not representative of the full ED patient population: its members are younger, more digitally active, and more likely to experience language barriers than the average Danish emergency patient. The dataset amplifies certain themes and almost certainly underrepresents others, particularly the experience of elderly monolingual Danish patients with cognitive impairment. The thematic analysis should be understood as corroborating rather than establishing the significance of these themes.

A further limitation is source diversity. Although two independent threads were analysed, both were drawn from the same platform (Facebook), and each was captured at a single point in time in a group with evident moderation tendencies. The corroborating Aarhus thread partially addresses the single-group limitation, but both groups skew toward international residents who are digitally active. Future research using this methodological approach should seek multi-platform corpora, ideally including Danish-language sources, alongside explicit archival strategies that account for moderation effects.

### 6.3 The Prototype in the Broader Digital Health Landscape

The ED Adaptive Interface sits within a growing landscape of patient-facing digital tools deployed in clinical settings. Commercial proprietary ED tablet solutions have been implemented in larger European hospital groups, typically with single-language interfaces, standard VAS pain scales, and call-nurse buttons without categorical differentiation. The absence of the features documented in this paper from commercial implementations suggests either that the problem has not been identified or that design priority has not been placed on the most constrained users. The prototype demonstrates that building this kind of system is neither technically complex nor expensive: it required no specialist equipment, no proprietary software, and no institutional budget. The primary investment was in design thinking.

### 6.4 On Convergent Validity

The convergence between the design rationale developed independently from the literature and the themes emerging independently from the social media corpus is meaningful evidence. The Workflow Tracker was not designed because of the "Foreigners in Denmark" thread — it was designed because alarm fatigue research indicated it would reduce nursing interruptions. The thread then independently produced 31 entries describing exactly the kind of navigation and information confusion the Workflow Tracker is designed to address. This convergence does not prove efficacy, but it does suggest that the problems being addressed are real, widely experienced, and not artefacts of the specific design context.

The most striking convergent finding is Theme 1’s structural pattern: the most highly-engaged comments in the thread were factually incorrect about whether the original poster had appropriate grounds for her attendance. This is a finding about the information environment in which patients experience and discuss emergency care. The Referral Pathway Display and Why Am I Waiting? module change the information available to the patient — a modest intervention with potentially significant effects on patient anxiety, social sanction, and self-discharge rates.

## 7. Limitations

Several limitations of this work must be stated clearly. First and most significantly, the ED Adaptive Interface has not been tested with real patients or clinical staff. All design decisions, however well-grounded in published literature and social evidence, remain unvalidated in clinical practice. The prototype is ready for a structured pilot; it is not yet evidence-based in the clinical sense.

Second, the prototype’s current architecture lacks server-side data persistence, meaning that symptom logs, escalation alerts, and pain scores captured during a patient visit are not transmitted to or stored in any clinical information system. This limits the clinical utility of the Symptom Progression Log and Condition Worsening Escalation features in real deployment, pending the PMS integration described in Section 9.

Third, both social media corpora have the limitations described in Section 6.2: demographically skewed toward digitally active international residents, Facebook-platform only, and subject to group moderation effects. The thematic analyses represent qualitative corroboration from two independent sources, not quantitative evidence of design impact.

Fourth, language support, while broader than most comparable systems (seven languages in v5), remains substantially less than the full range of languages spoken by patients in Danish emergency departments. The prioritisation of Romanian, Polish, and Somali for v5 was informed by census data on non-Danish-speaking populations in the Region Hovedstaden catchment area, but necessarily excludes other communities.

Finally, WCAG 2.1 AA compliance of the high-contrast mode, while technically correct, has not been validated with patients who have specific colour vision deficiencies or complex visual impairments. WCAG compliance establishes a minimum standard; it does not guarantee usability for all members of the target population.

## 8. Conclusion

Forty-three per cent of emergency patients at Nordsjællands Hospital struggle with digital tools. This figure is the predictable consequence of deploying interfaces designed for baseline conditions in a setting where baseline conditions are routinely absent. The appropriate response is not to slow down digitalisation but to change how it is designed.

The ED Adaptive Interface demonstrates that it is technically and financially feasible to build a patient-facing digital tool that configures itself in under ten seconds to serve patients across the full spectrum of visual, motor, speech, and cognitive impairment — in seven languages, with zero infrastructure requirements, running on any existing hospital tablet. The impairment-first design philosophy that produced this prototype offers a transferable framework: begin with the most constrained user, solve their problem rigorously, and discover that you have also substantially improved the experience for everyone else.

The social media discourse analysis presented in this paper adds a form of evidence that formal patient satisfaction research rarely captures. Two independent social media corpora — 83 entries from the primary dataset and over 100 comments from the corroborating Aarhus thread — drawn from forums not designed for research, independently surfaced the same five categories of failure the prototype was built to address. That convergence is not proof of clinical efficacy; it is evidence that these problems are real, widely experienced, and structurally reproducible. Version 5 of the prototype, reported here for the first time, translates the findings of that analysis directly into implemented features.

The prototype is ready for a structured clinical pilot. The questions that remain — does it reduce nursing interruptions? Does it improve patient-reported anxiety? Does it increase pain score accuracy? — are answerable, and the roadmap for answering them is clear. The question is not whether a system like this can be built. It has been. The question is whether the institutions that need it most are ready to move.

> *"We don’t ask the patient to learn our technology; our technology learns the patient."*

## 9. Future Work

### 9.1 Clinical Pilot Design

The immediate next step is a structured clinical pilot at Nordsjællands Hospital. Phase 1 would involve usability testing with five to ten ED nurses and a comparable number of patients across two dedicated testing days, using the System Usability Scale (SUS) [16] as the primary quantitative instrument. Task completion rates — nurse setup time, first successful pain rating, help request submission — would provide supplementary evidence. Semi-structured interviews with nursing staff would surface workflow integration concerns that desk-based prototyping cannot anticipate.

Phase 2 would involve a four-week ward pilot on existing tablet hardware in the Emergency Department, without modification to the ward’s IT infrastructure. Patient-reported outcomes (anxiety, perceived communication quality, understanding of care process) would be collected at discharge using brief validated instruments. Nursing workload indicators — specifically the frequency of call-button activations and status-update requests — would be tracked against baseline data to provide a preliminary estimate of workflow impact.

Phase 3, contingent on Phase 2 findings, would address PMS integration for live workflow stage updates and export of patient-reported symptom data to nursing dashboards.

### 9.2 Full MinSP Integration

MinSP is Denmark’s national citizen health record platform. If a patient’s accessibility profile, language preference, chronic condition history, and relevant medical context could be pre-populated from MinSP at arrival — triggered by scanning the patient’s yellow card (Sundhedskort) with an NFC reader or barcode scanner — the nurse setup step would be reduced to a single confirmation. This is a significant technical undertaking involving API access agreements with the Danish Health Data Authority, GDPR-compliant authentication protocols, and integration testing with the hospital’s existing PMS. The MinSP simulation feature in v5 demonstrates this concept and serves as a stakeholder communication tool for the integration roadmap.

### 9.3 Additional Language Support

Version 5 supports seven languages. The most common non-Danish, non-English languages spoken by patients in the Region Hovedstaden catchment area include Urdu, Dari/Farsi, Tigrinya, Vietnamese, and several West African languages not currently supported. Expanding language coverage to the top fifteen non-Danish languages in the region is a realistic near-term goal; the translation framework in the current prototype is structured to accept new language objects without requiring changes to the interface logic. Any new language additions should be verified by bilingual clinicians or professional medical translators before deployment.

### 9.4 Hospital PMS Integration for Live Workflow Updates

Currently, workflow stage transitions are triggered manually via the Staff Simulation Mode. In a deployed system, these transitions should be triggered automatically by the hospital’s PMS — specifically by status updates already being logged in the clinical workflow (blood test ordered, results received, doctor assigned). PMS integration represents the single highest-impact technical upgrade to the deployed system, allowing the workflow tracker and care notifications to function continuously without imposing any additional staff burden.

### 9.5 Paediatric and Companion-Assisted Modes

The current prototype is designed for adult patients. Paediatric patients present distinct interface requirements: simpler language, larger visual elements, and a companion-mediated configuration where a parent or guardian interacts with the system on the child’s behalf. A companion-assisted mode — where a family member or carer is explicitly designated as the interface operator — would also support patients with severe cognitive impairment for whom nurse setup cannot anticipate all interaction needs.

## Data Availability

The anonymised social media discourse dataset is archived by the corresponding author and is available to reviewers upon reasonable request. The prototype (ED Adaptive Interface v5) is a self-contained HTML file available from the corresponding author upon request. No other primary data were generated.

## Acknowledgements

The initial solution concept was developed during the OpenInnovation 2026 Health challenge (February 2026), a cross-university student innovation sprint co-organised by DTU Skylab, Copenhagen School of Entrepreneurship (CBS), and KU Actory (University of Copenhagen), with Nordsjællands Hospital Emergency Department as case partner. The prototype, research methodology, social media analysis, and all work reported in this paper were developed independently by the author after the challenge period (February–April 2026). The author thanks Professor Thomas Andersen Schmidt and Jens-Peter Baatz Kristensen (Head of Innovation, Nordsjællands Hospital) for presenting the challenge brief and for their engagement with all participating teams. The social media discourse dataset was collected and anonymised by the author independently; no institutional data collection infrastructure was involved. Manuscript preparation assistance was provided by Atashi Irtiza, MBBS, DCO, Bangladesh Medical University. Generative AI tools were used to assist with drafting, writing, and prototype code development during the preparation of this manuscript. All scientific content, data collection, analysis, and conclusions are the sole responsibility of the authors.

## Data Ethics Statement

The secondary evidence reported in Section 5 consists of publicly accessible social media content collected from a Facebook group ("Foreigners in Denmark") on 14 March 2026. The data collection and analysis were conducted in accordance with the following ethical provisions:

- Public accessibility: The group and the thread analysed were publicly accessible at the time of collection. No private group membership was required to view the content, and no authentication credentials were used to access it.
- No participant contact: No commenter was contacted before, during, or after the analysis. No consent was sought from individual participants, consistent with GDPR Article 89 provisions for research processing of publicly available data.
- Full anonymisation: All identifiable information — names, usernames, profile images, and any biographical details — was removed from the dataset and replaced with non-identifiable reference codes (C01–C81 for commenters, OP for the original poster) prior to any coding or analysis. No identifiable information appears in this paper or in the research archive.
- No participant harm: The analysis did not involve any interaction with participants and produced no information that could affect any individual’s healthcare, employment, legal status, or reputation.
- GDPR compliance: The processing of this data for research purposes is conducted under GDPR Article 89, which permits the use of personal data for scientific research where appropriate safeguards — including anonymisation — are applied. No institutional ethics approval is required for retrospective anonymised analysis of publicly posted content under Danish research practice for this class of data.
- Data archival: The anonymised dataset is archived by the author and is available to reviewers upon reasonable request. The original non-anonymised data has been permanently deleted from all author systems following the anonymisation process.

A second sample was gathered in April 2026 in a Facebook group that serves the international community in the Aarhus area of Denmark. All the above-described anonymisation, non-contact, and GDPR Article 89 provisions are applicable to this dataset. All identifiers such as names, usernames, profile pictures and any location or biographical information were deleted and substituted with non-identifiable reference codes before analysis. None of the participants were contacted. All author systems have been permanently destroyed with all the non-anonymised information. The researchers wrote this Data Ethics Statement, which is not an official institutional ethics board decision. The authors verify that this study design did not need any institutional ethics review according to the existing regulations.

